# CNBP, REL, and BHLHE40 variants are associated with IL-12 and IL-10 responses and tuberculosis risk

**DOI:** 10.1101/2021.03.03.21252797

**Authors:** Javeed A. Shah, Alex J. Warr, Andrew D. Graustein, Aparajita Saha, Sarah J. Dunstan, Nguyen T.T. Thuong, Guy E. Thwaites, Maxine Caws, Phan V.K. Thai, Nguyen D. Bang, Tran T.H. Chau, Felicia K. Nguyen, Carlo A. Hernandez, Madison A. Jones, Christopher M. Sassetti, Katherine A. Fitzgerald, Munyaradzi Musvosvi, Anele Gela, Willem A. Hanekom, Mark Hatherill, Thomas J. Scriba, Thomas R. Hawn

**Affiliations:** University of Washington, Seattle, USA; VA Puget Sound Health Care System. Seattle, USA; University of Melbourne, Melbourne, Australia; Oxford University Clinical Research Unit, Ho Chi Minh City, Vietnam; Centre for Tropical Medicine and Global Health, Nuffield Department of Medicine, University of Oxford, Oxford, UK; Liverpool School of Tropical Medicine, UK; Pham Ngoc Thach Hospital, Ho Chi Minh City, Vietnam; University of Massachusetts, Worchester, MA; South African Tuberculosis Vaccine Initiative, Cape Town, South Africa

**Keywords:** CNBP, REL, BHLHE40, dendritic cells, genetics, *M. tuberculosis*

## Abstract

**Rationale:** The major human genes regulating *M. tuberculosis* (Mtb)-induced immune responses and tuberculosis (TB) susceptibility are poorly understood. Although IL-12 and IL-10 are critical for TB pathogenesis, the genetic factors that regulate their expression are unknown. CNBP, REL, and BHLHE40 are master regulators of IL-12 and IL-10 signaling.

**Objectives:** To determine whether common human genetic variation in CNBP, REL and BHLHE40 is associated with IL-12 and IL-10 expression, adaptive immune responses to mycobacteria, and susceptibility to TB.

**Methods and Main Measurements:** We characterized the association between common variants in CNBP, REL, and *BHLHE40* and innate immune responses in dendritic cells and monocyte-derived macrophages (MDM), BCG-specific T cell responses, and susceptibility to pediatric and adult TB.

**Results:** SNP BHLHE40 rs4496464 was associated with increased BHLHE40 expression in MDMs and increased IL-10 from both peripheral blood dendritic cells and MDMs after LPS and TB whole cell lysate stimulation. SNP BHLHE40 rs11130215, in linkage disequilibrium with rs4496464, was associated with increased BCG-specific IL2+CD4+ T cell responses and decreased risk for pediatric TB in South Africa. SNPs REL rs842634 and CNBP rs11709852 were associated with increased IL-12 production from dendritic cells, and SNP REL rs842618, in linkage disequilibrium with rs842634, was associated with increased risk for TB meningitis.

**Conclusions:** Genetic variation in CNBP, REL, and BHLHE40 is associated with IL-12 and IL-10 cytokine response and TB clinical outcomes. Common human genetic regulation of well-defined intermediate cellular traits provides insights into mechanisms of TB pathogenesis.

## Introduction

Tuberculosis (TB) is a leading cause of death from infection worldwide. The current BCG vaccine remains the only approved vaccine against TB despite its partial and variable effects across populations (1). Vaccine efforts are hampered by a lack of understanding of the immune correlates of protection (2). Understanding the factors required to induce effective, long lasting immunity to infections may provide tools to improve TB vaccines.

Twin, Mendelian, linkage, genome-wide association, and candidate gene studies suggest that genetic factors influence susceptibility to TB (3, 4). Multiple clinical TB phenotypes show a high degree of heritability, including host susceptibility to pulmonary TB (5–10), TB meningitis (11, 12), and latent TB infection (13–17). However, the major genes regulating TB susceptibility have not yet been identified with consistent results across multiple populations, possibly due to heterogeneous clinical phenotypes and lack of mechanistic correlation of genetic variants with immunophenotypes (3). To overcome these obstacles, we evaluated LPS and Mtb whole cell lysate (TBWCL)-induced cytokine responses from immune cells, followed by clinical correlation, to improve the power and mechanistic insight of genetic studies.

Common genetic variation influences the cellular innate immune response to *Mycobacterium tuberculosis* (Mtb). Multiple studies demonstrate the impact of genetic variation on innate immune cellular distribution and cytokine responses (18–21). Quantitative trait loci (QTL) of gene expression demonstrate immune cell-specific effects (22). Recent advances permit the evaluation of innate immune cytokine responses from rare cell populations (23, 24). Variants that influence functional responses in immune cells of interest represent attractive secondary traits which can be correlated with TB susceptibility and these correlations may provide insight into genetic mechanisms of disease susceptibility (25).

Dendritic cells (DCs) present antigen to T cells via MHC Class I and II, co-stimulate them with CD40 and CD80, and influence T cell differentiation by producing cytokines like IL-12p70, IL-10, and IL-23, to induce T cell differentiation (26). DCs are essential for mycobacterial immunity (15, 27) and common genetic variants that influence DC migration are also associated with TB susceptibility (7). IL-10 and IL-12 are particularly important for T cell function in Mtb infection. Individuals with Mendelian deficiencies in IL-12 signaling rapidly develop serious, disseminated mycobacterial infections (28, 29). However, the effect of common genetic variation on physiologic levels of IL-10 and IL-12, and the influence of these cytokines on BCG-specific T cell responses and TB outcomes in humans is not known. After inflammatory stimulation, the transcription factor CNBP and its binding partner c-REL translocate to the nucleus and induce IL12B transcription, which encodes the IL-12p35 protein subunit (30, 31). Likewise, IL-10 production influences Mtb immune responses, as it diminishes T cell activation, enhances regulatory T cell activity, and may be responsible for delayed T cell priming observed in the initial Mtb immune response (32, 33). In mice, the transcription factor BHLHE40 controls IL-10 production from both myeloid and lymphoid cells, with contribution from CNBP (30, 31, 34). The role of these genes and their genetic variants in human regulation of T cell responses is unknown. In this study, we investigated whether common human genetic variation in the transcription factors CNBP, REL, and BHLHE40 were associated with DC cytokine responses, BCG-specific T cell responses and TB susceptibility.

## Materials and Methods

### Ethics Statement

Approval for human study protocols was obtained from the institutional review boards at local sites and at the University of Washington School of Medicine (Seattle, WA). The South African study included written informed consent from the parent or legal guardian of the participant and approval by the University of Cape Town Research Ethics Committee. Written informed consent was received from all participants before inclusion in the study. For genetic studies in Vietnam, approval for human study protocols was obtained from the human subjects review boards at the University of Washington School of Medicine, the Hospital for Tropical Diseases, Pham Ngoc Thach Hospital, Hung Vuong Hospital, and the Oxford Tropical Research Ethics Committee. Written informed consent was obtained from patients or their relatives if the patient could not provide consent.

### Study Participants

Study participants in the Seattle cohort were volunteers self-described as healthy without history of recurrent serious infections. 52% of individuals were female, and 48% were male. The ethnic composition of this study group was 69% White, 19% Asian, 2% Black or African American, and 2% Latinx. Average age of study participants was 39, with interquartile range of 29 – 46 at the time of their enrollment.

South African study participants were enrolled at the South African Tuberculosis Vaccine Initiative field site in Worcester, South Africa, near Cape Town as part of a larger study on BCG vaccination with 11,680 infants (35, 36). This area has one of the highest rates of TB in the world with an incidence of 3% among children less than 3 years of age in the study population (35, 36). A nested genetics case-control study was performed with identification of cases and controls during a 2-year prospective observation period after vaccination at birth. The criteria for detection of TB cases have been described previously and are summarized in the online supplement (37).

Study subjects from the Vietnam cohort were described previously and are summarized here and in detail in the online supplement (12). Subjects with tuberculous meningitis were recruited from two centers in Ho Chi Minh City, Vietnam: Pham Ngoc Thach (PNT) Hospital for Tuberculosis and the Hospital for Tropical Diseases (HTD). Subjects with pulmonary TB were recruited from a network of district TB control units within Ho Chi Minh City that provide directly observed therapy to TB patients. In addition, pulmonary TB subjects enrolled were recruited from PNT hospital from 2006 through 2008. Vietnamese population controls were otherwise healthy adults with primary angle closure glaucoma which have been previously described (38). All case and control participants were unrelated and greater than 95% were of the Vietnamese Kinh ethnicity. Previous genetic studies of this population indicate minimal population substructure (12, 39).

All statistical analyses are described in the online supplement and were performed using Stata 14.1 and Prism 8.0 software. The remainder of all experimental procedures are described in detail in the online supplement.

## Results

### Single cell analysis of cytokine production in peripheral blood DCs

To evaluate genetic regulation of IL-10 and IL-12 production from healthy human donors, we used flow cytometry to measure the proportion of peripheral blood MHC-II+CD11c+ DCs producing IL-10 and IL-12 after stimulation of whole blood with LPS or TB whole cell lysate (TBWCL; Figure 1A). LPS (10 ng/ml) and TBWCL (50 µg/ml) both strongly induced IL-12 (Figure 1B) and IL-10 (Figure 1C) from DCs 24 hours after stimulation. We also measured cytokine responses to LPS (10 ng/ml) and live BCG (10^6^ CFU/ml) 6 hours after stimulation (Figure 1D). We found that LPS and BCG induced IL-12 6 hours after stimulation in CD11c+ DCs. However, we did not detect IL-10 above background levels from DCs after 6 hours of stimulation (data not shown).

**Figure 1.**
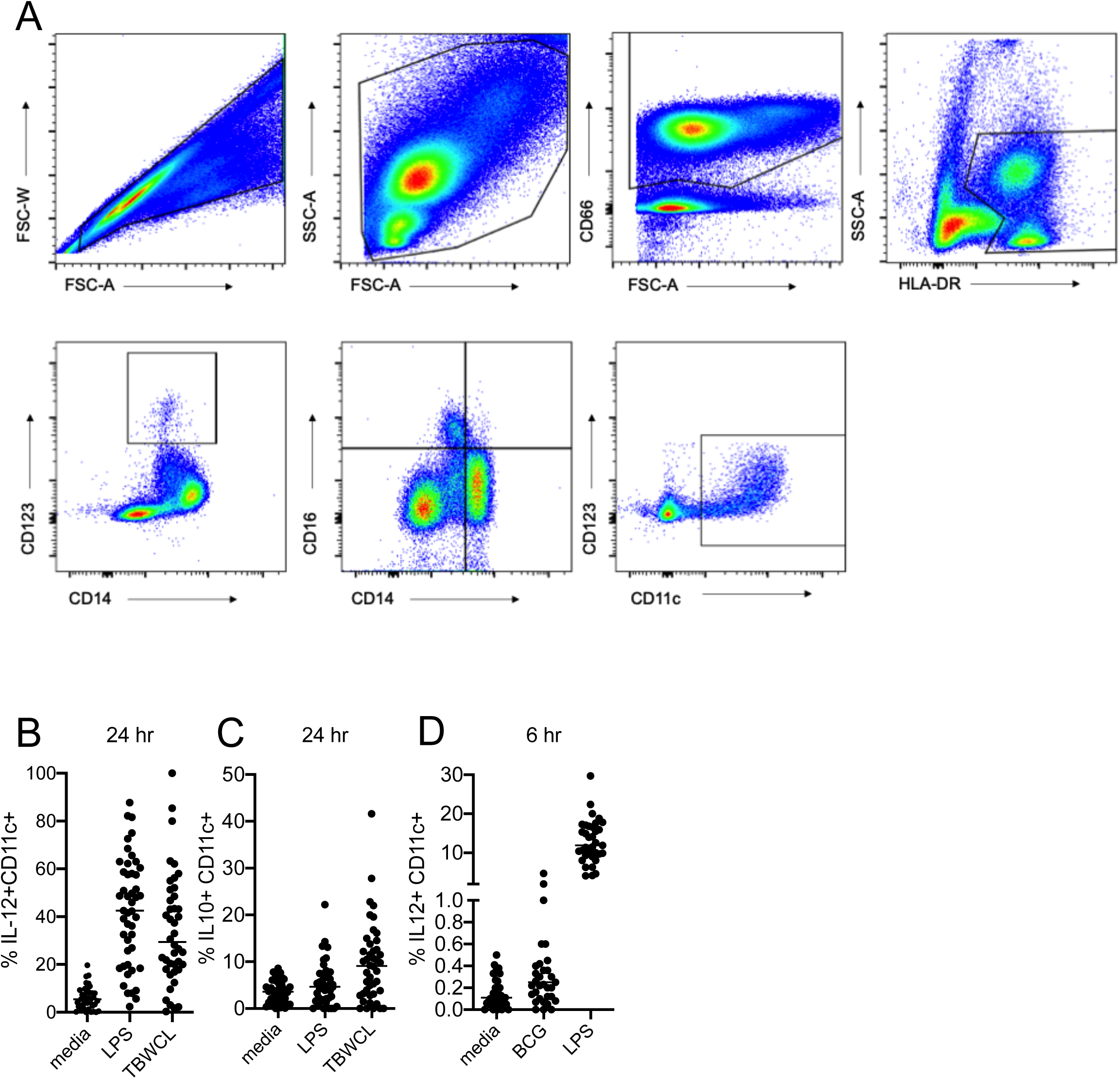
IL-10 and IL-12 responses in peripheral blood DCs in whole blood stimulation assay. Peripheral whole blood was obtained from healthy volunteers and stimulated with either negative control or immune stimuli followed by BFA and monensin 2 hours afterward. Afterward cells were fixed and frozen. At the time of staining, samples were thawed in large batches to minimize batch effects. A) Gating strategy. From *left* to *right*, singlets were selected, then leukocytes. CD66+ cells were gated out, and the HLA-DR+ population selected. CD14- and CD16- and CD11c+ cell population was selected and the proportion of cytokine positive cells were measured as compared to total number of HLA-DR+CD11c+ DCs. B) Proportion of IL-12+CD11c+ DCs after media control, LPS (10^6^ ng/ml), or Mtb whole cell lysate (TBWCL; 50 µg/ml) stimulation for 24 hours. C) Proportion of IL-10+CD11C+ DCs after media, LPS, or TBWCL for 24 hours. D) Proportion of IL-12+CD11c+ DCs after media, LPS, or live BCG (10^6^ CFU) stimulation for 6 hours. Bars demonstrate median values. Data provided are not corrected for background cytokine positivity. Dots represent individual values. N = 46.

### Discovery analysis of genetic associations with IL-12 responses to LPS and TBWCL

We next examined whether candidate gene variants were associated with LPS or TB whole cell lysate- (TBWCL) induced IL-12 in DCs. We interrogated 4 haplotype-tagging SNPs from CNBP, 6 from REL, and 19 from BHLHE40 in a local cohort of healthy volunteers (Figure E1). REL SNP rs842634 was associated with increased IL-12 after TBWCL and LPS stimulation (Figure 2A; p = 0.044, generalized linear model (GLM), Figure 2B; p = 0.037). CNBP SNP rs11709852 was associated with increased IL-12 production after TBWCL stimulation, but not LPS stimulation (Figure 2C; p = 0.003; Figure 2D, p = 0.48). No SNPs from BHLHE40 were associated with IL-12 (Table E2).

**Figure 2.**
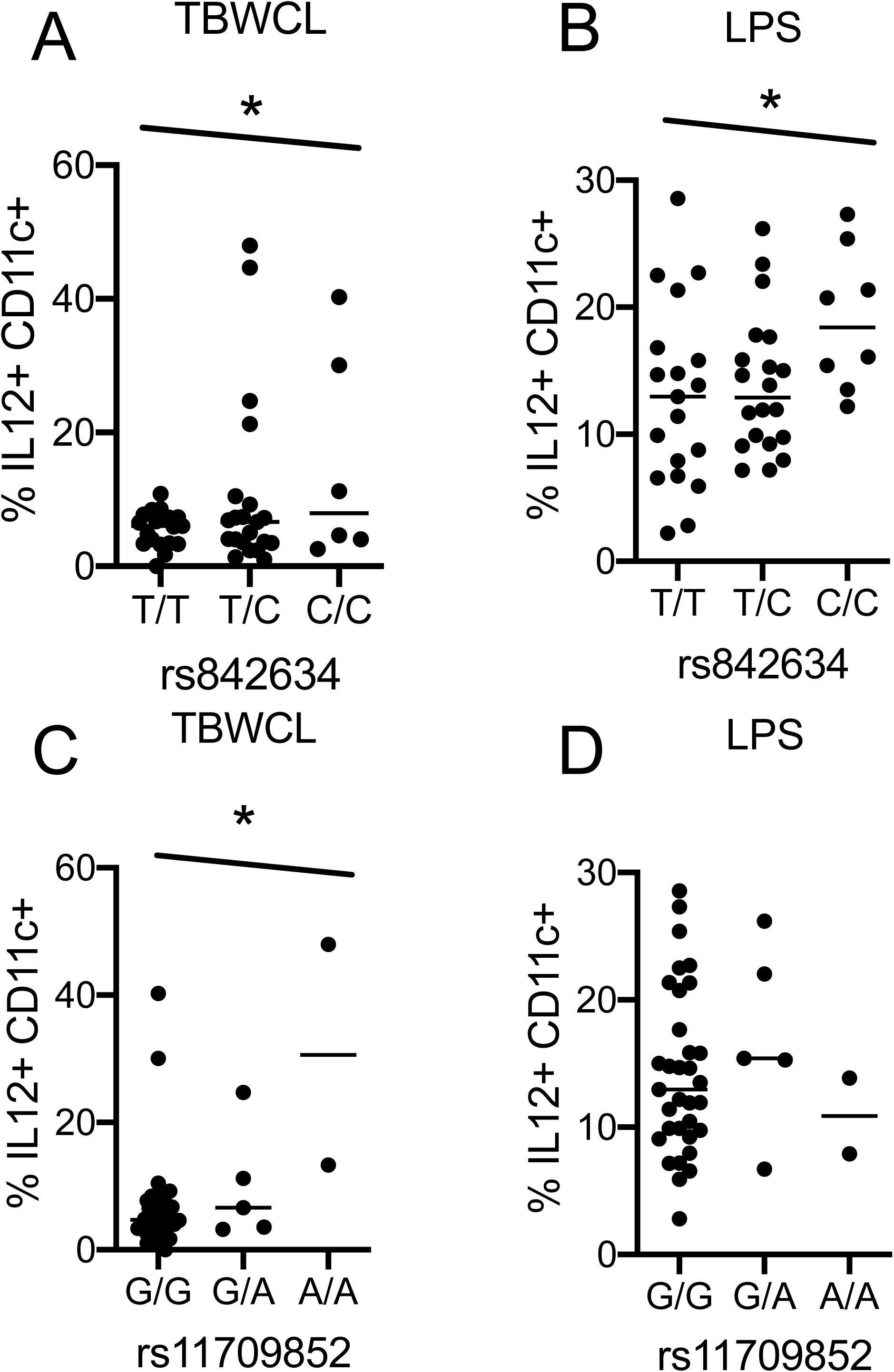
REL SNP rs842634 and CNBP SNP rs11798052 are associated with IL-12 production after TBWCL stimulation of peripheral blood DCs for 24 hours. A-B) Proportion of CD11c+ DCs producing IL-12 after A) Mtb whole cell lysate (TBWCL; 50 µg/ml) stimulation or B) LPS (10 ng/ml) stimulation for 24 hours. Data are stratified by rs842634 genotype; N = 19 T/T, 21 T/C, and 7 C/C. C-D) Proportion of CD11c+ DCs producing IL-12 after C) TBWCL or D) LPS stimulation for 24 hours. Data are stratified by rs11798052 genotype; N = 34 G/G, 5 G/A, and 2 A/A. All data presented in this figure and afterward represent background-corrected values (proportion of cytokine-producing cells after ligand stimulation – proportion of cytokine-producing cells after media control stimulation). * p < 0.05; statistical significance determined by generalized linear model.

### CNBP and REL variants are associated with LPS and BCG-induced IL-12 secretion after 6 hour stimulation in an independent dataset

We evaluated the association of our candidate SNPs in a second, independent cohort with whole blood stimulated with BCG (10^6^ CFU/ml) or LPS (10 ng/ml) for 6 hours, followed by measurement of cytokine responses, as described above. REL SNP rs842634 was associated with increased IL-12 after BCG infection (Figure 3A; p = 0.046, generalized linear model) and LPS stimulation (Figure 3B; p = 0.024). CNBP SNP rs11709852 was associated with a trend toward increased IL-12 after BCG stimulation (Figure 3C; p = 0.078, Mann-Whitney U-test), and was also associated with increased IL-12 after LPS stimulation early in infection Figure 3D; p = 0.014, Mann-Whitney test).

**Figure 3.**
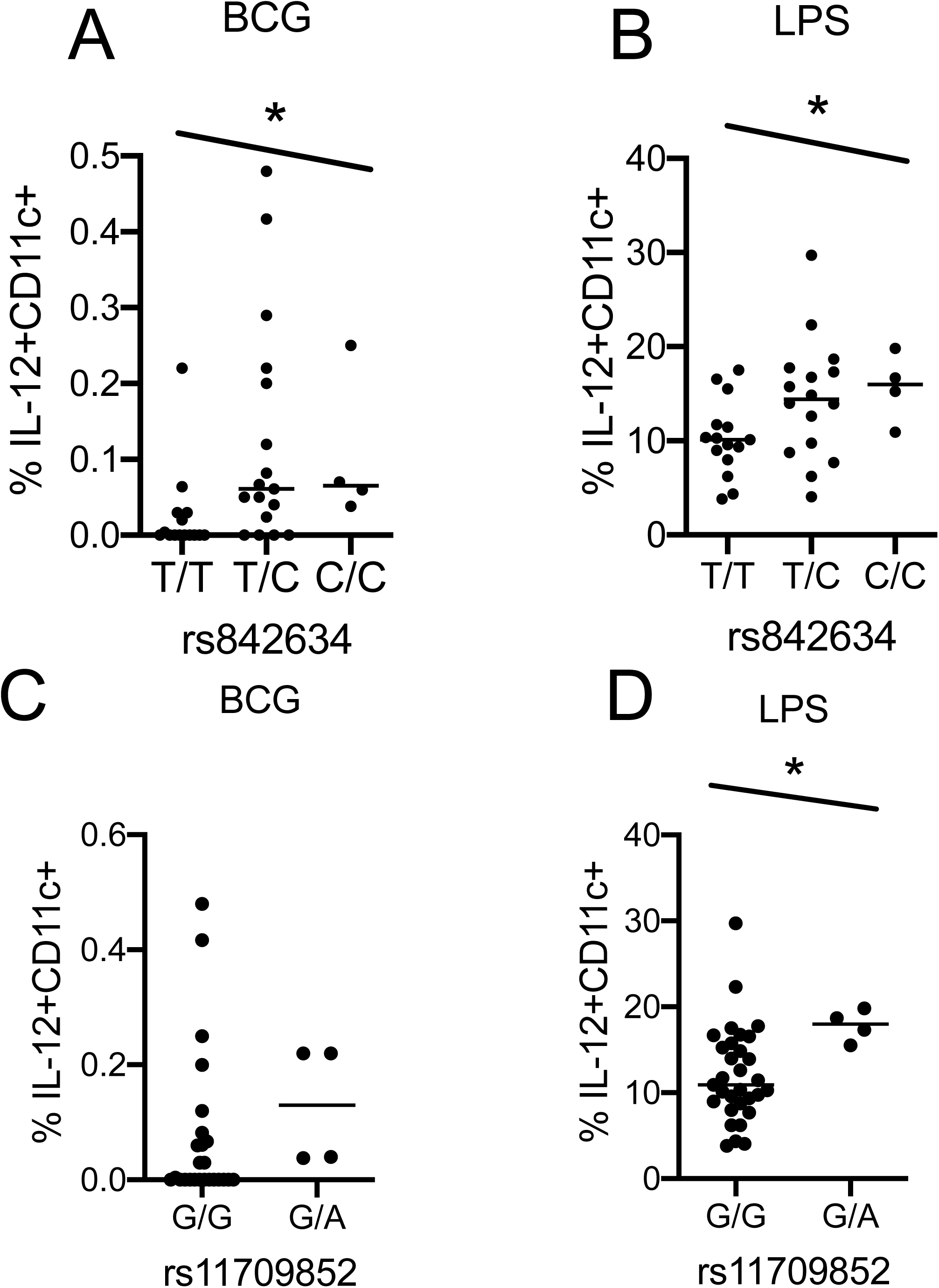
REL SNP rs842634 is associated with IL-12 production in peripheral blood DCs after 6 hours of BCG or LPS stimulation. A-B) Proportion of CD11c+ DCs producing IL-12 after A) live BCG stimulation (10 CFU) or B) LPS (10^6^ ng/ml) stimulation for 6 hours. Data are stratified by rs842634 genotype; N = 15 T/T, 16 T/C, and 4 C/C. C-D) Proportion of CD11c+ DCs producing IL-12 after C) live BCG stimulation or D) LPS stimulation for 6 hours. Data are stratified by rs11798052 genotype; N = 31 G/G, 5 G/A. * p < 0.05; ** p < 0.01, *** p < 0.001; statistical significance determined by generalized linear model for A-B and Mann-Whitney U-test for C-D.

### BHLHE40 SNP rs4496464 is associated with IL-10 secretion from DCs

Next, we evaluated for associations between genetic variants in CNBP, REL, and BHLHE40 with IL-10 production from DCs. BHLHE40 SNP rs4496464 was associated with increased IL-10 production after TBWCL stimulation (Figure 4A; p = 0.005, generalized linear model). In contrast, rs4496464 was not associated with IL-10 after LPS stimulation (Figure 4B, p = 0.18). No CNBP or REL SNPs, including rs11709852 and rs842634, were associated with IL-10 expression after TBWCL or LPS stimulation. (Figure 4C – F). BHLHE40 SNP rs4496464 was not associated with IL-12 expression after stimulation with either TBWCL or LPS (Figure 4G and Figure 4H).

**Figure 4.**
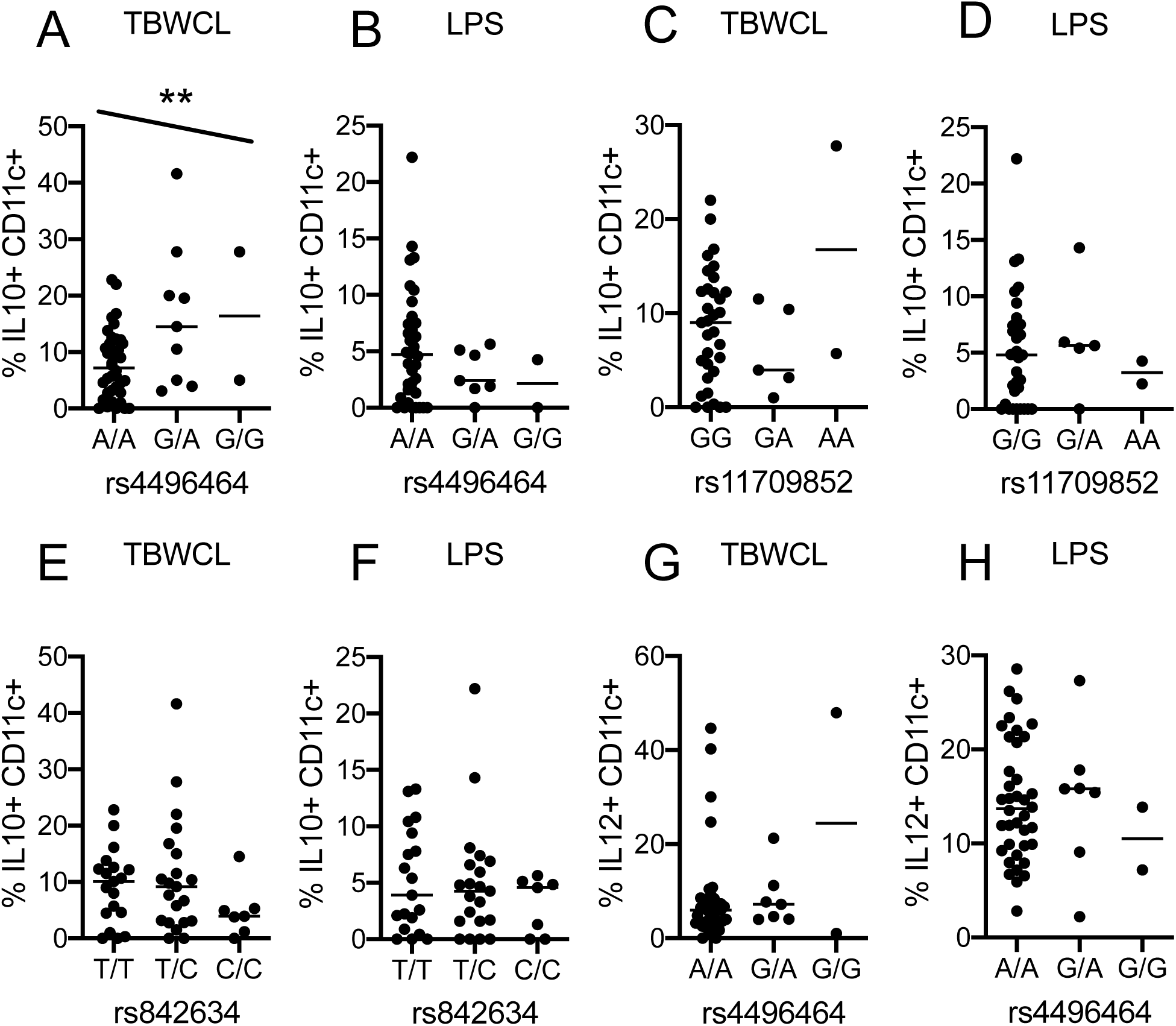
BHLHE40 SNP rs4496464 is associated with IL-10 production from peripheral blood DCs after Mtb whole cell lysate stimulation. A-B) Proportion of CD11c+ DCs producing IL-10 after A) Mtb whole cell lysate (TBWCL; 50 µg/ml) or B) LPS (10 ng/ml) stimulation for 24 hours. Data are stratified by rs4496494 genotype; N = 40 A/A, 7 G/A and 2 G/G. C-D) Proportion of CD11c+ DCs producing IL-10 after C) LPS or D) TBWCL stimulation for 24 hours. Data are stratified by rs11798052 genotype; N = 33 G/G, 5 G/A, and 2 A/A. E-F) Proportion of CD11c+ DCs producing IL-10 after E) LPS or F) TBWCL stimulation for 24 hours. Data are stratified by rs842634 genotype; n = 19 T/T genotype, 21 T/C genotype, and 7 C/C genotype. G-H) Proportion of CD11c+ DCs producing IL-12 after E) TBWCL or F) LPS stimulation for 24 hours. Data are stratified by rs4496494 genotype. N = 38 A/A, 7 G/A, 2 G/G. * p < 0.05; ** p < 0.01, *** p < 0.001; generalized linear model.

### Rs4496464 is associated with BHLHE40 mRNA expression in monocyte-derived macrophages

We evaluated whether rs4496464 genotypes were associated with BHLHE40 mRNA expression in peripheral blood monocyte-derived macrophages (MDM) from healthy donors. The uncommon G allele of rs4496464 was associated with increased BHLHE40 in unstimulated monocytes using a dominant model of inheritance (Figure 5; p = 0.026, A/A vs (G/A + G/G), Mann-Whitney U-test). No other BHLHE40 SNPs were associated with expression. There was no association in LPS stimulated monocytes. CNBP and REL variants were not associated with their respective transcripts (data not shown).

**Figure 5.**
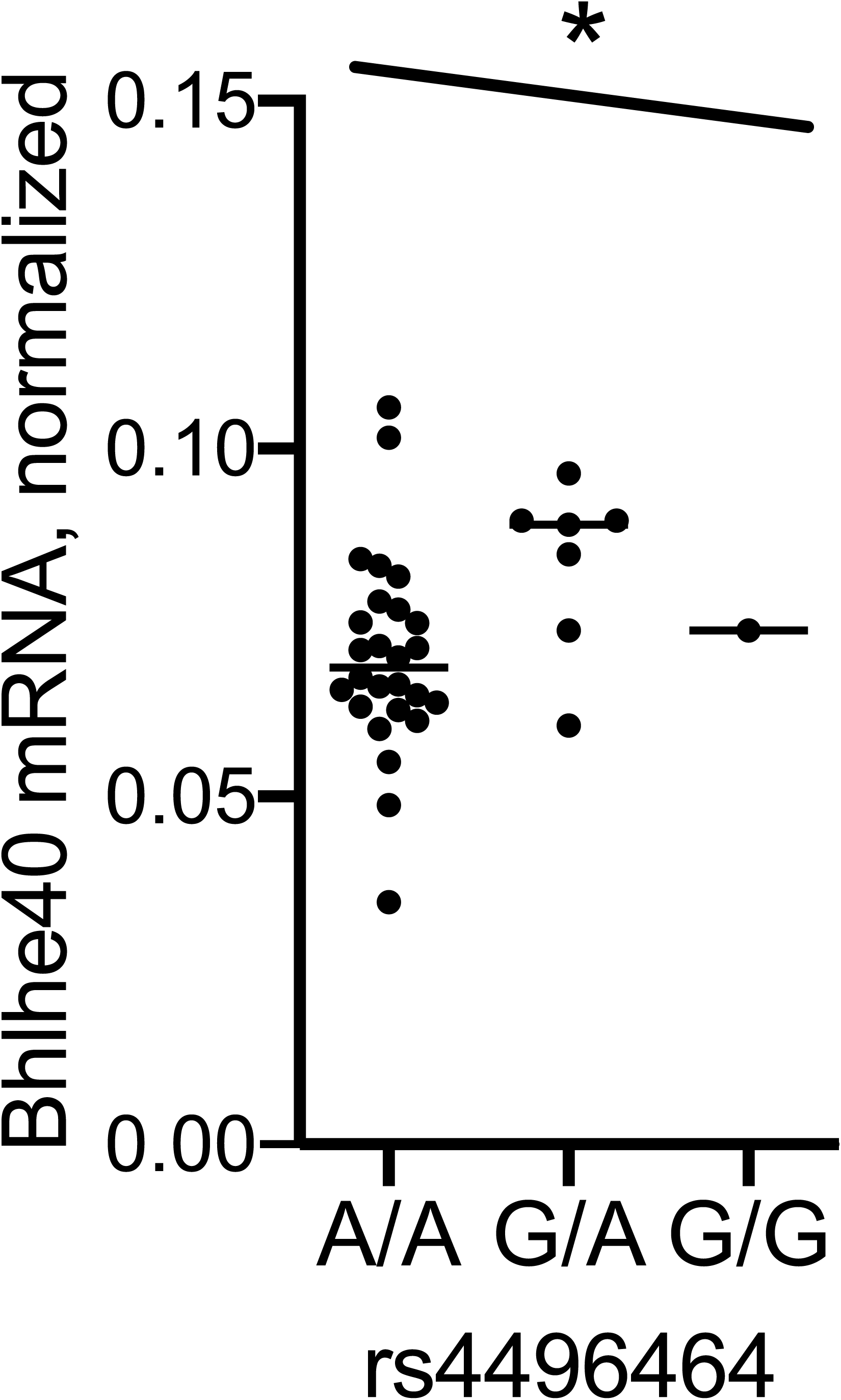
BHLHE40 SNP rs4496464 is associated with increased BHLHE40 mRNA expression in monocyte-derived macrophages. BHLHE40 mRNA expression, normalized to GAPDH expression, was measured from RNA extracted from MDMs isolated from healthy volunteers and stratified by rs4496464; n = 26 A/A, 7 G/A, and 1 G/G. * p < 0.05; dominant genetic model.

### Rs4496464 is associated with IL-10 production in LPS and TBWCL stimulated monocyte-derived macrophages

To validate our association between rs496464 and IL-10 expression in DCs, we measured IL-10 secreted from monocyte-derived macrophages (MDMs) stimulated with either LPS (50 ng/ml) or TBWCL (25 µg/ml) overnight (Figure 6A, n = 26). The rs4496464 G allele was associated with increased IL-10 after LPS stimulation (Figure 6B, p = 0.01, generalized linear model). SNP rs4496464 was also associated with increased IL-10 after TBWCL (Figure 6C, p = 0.005, generalized linear model). SNP rs4496464 was not associated with TNF secretion after either LPS (Figure 6D) or TBWCL stimulation (Figure 6E), which suggests that variation in BHLHE40 is associated with IL-10 production specifically, over proinflammatory cytokine responses.

**Figure 6.**
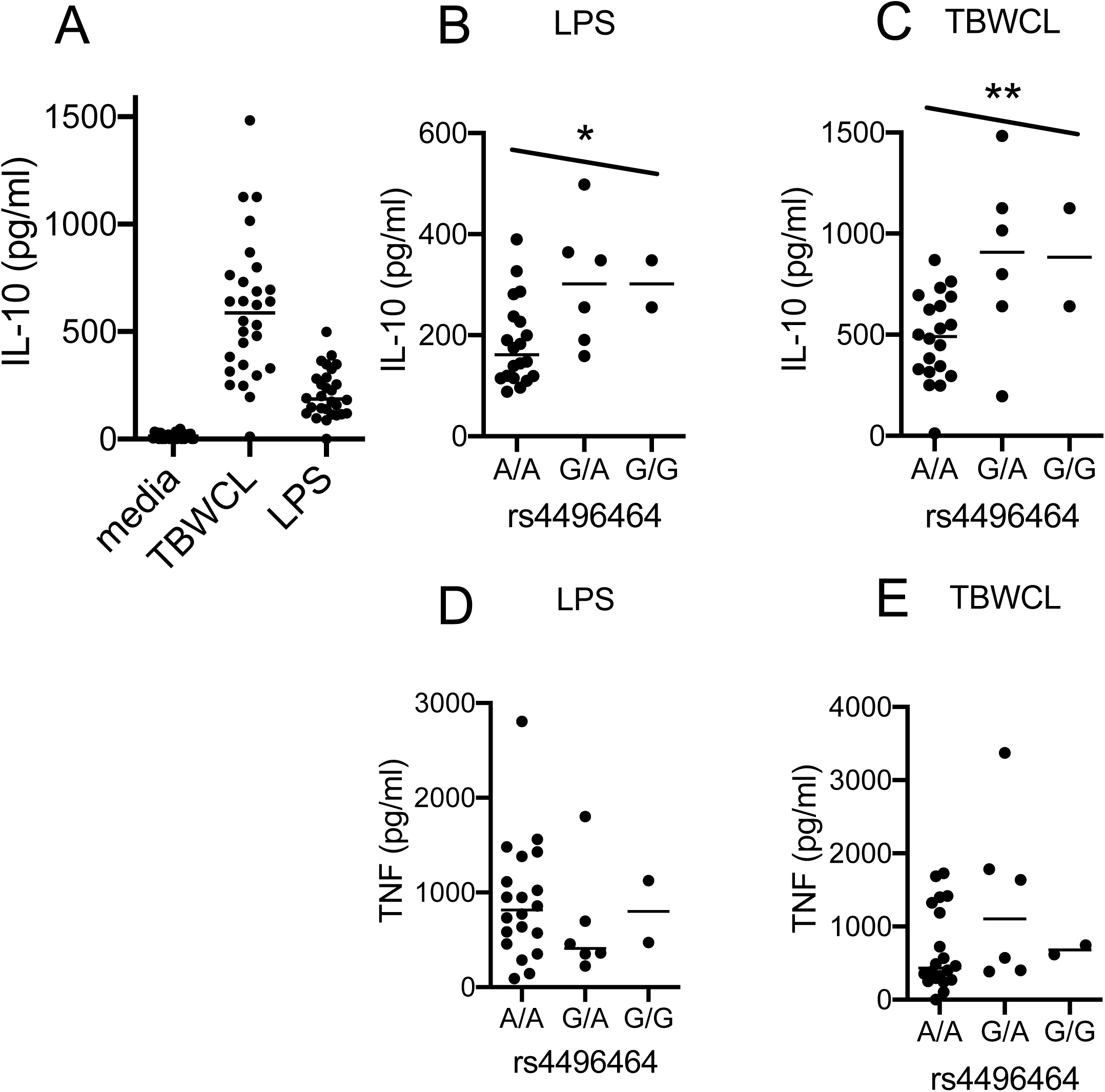
BHLHE40 SNP rs4496464 is associated with IL-10 production from monocyte-derived macrophages. Peripheral blood monocytes were differentiated into macrophages by M-CSF for 5 days, then stimulated with either LPS (50 ng/ml) or Mtb whole cell lysate (TBWCL; 25 µg/ml). A) Overall IL-10 cytokine concentrations from cellular supernatants MDMs after 24 hours of stimulation. B-C) Concentration of IL-10 in cellular supernatants after B) LPS stimulation or C) TBWCL stimulation for 24 hours, stratified by rs4496494 genotype. N = 20 A/A, 6 G/A, 2 G/G. D-E) Concentration of TNF in cellular supernatants after D) LPS stimulation or E) TBWCL stimulation for 24 hours and stratified by rs4496464. * P < 0.05, ** P < 0.01, *** P < 0.001; generalized linear model.

### A genetic marker for REL rs842634 is associated with an increased risk for TB meningitis

Our data suggests that rs842634 and rs11709852 are associated with increased IL-12 in DCs and rs4496464 is associated with increased IL-10 production from peripheral blood monocytes and DCs in our local population. We hypothesized that these polymorphisms are associated with susceptibility to TB due to their influence on these critical immune phenotypes. Within a large genome wide association study comparing Vietnamese individuals with adult pulmonary TB (PTB; n =1598) or TB meningitis (TBM; N = 407) with control subjects (N = 1139), we evaluated if SNPs in CNBP, REL, and BHLHE40 were associated with adult PTB or TBM and in LD with our SNPs of interest (Figure E2). Although REL rs842634 was not associated with TBM, it was in moderate to high LD with rs842618 in the Seattle cohort (R^2^ 0.69, D’ 1.0) as well as in the Vietnamese population (R^2^ 0.39, D’ 1.0). The minor allele of REL SNP rs842618 was associated with an increased risk for TBM (p = 0.03; OR 1.27, allelic model, Table 1 and Table E3). These data best fit a dominant model (Table 1, p = 0.035, OR 1.32, 95% CI 1.02 – 1.73) No BHLHE40 or CNBP SNPs were associated with TBM, including rs4496464 and rs11709852. We did not identify any associations between SNPs in REL, CNBP, or BHLHE40 SNPs with PTB (Table E4). Together, these data suggest that a causal REL SNP linked to rs842634 and rs842618 is associated with both increased IL-12 production and increased risk of adult TBM in Vietnam.

**Table 1.** Association of REL SNPs with adult TB meningitis in Vietnam. Number of individuals with major homozygous (AA), heterozygous (Aa), and minor homozygous (aa) genotypes described. Total: total N in group after genotyping. Allelic p: p value in an allelic genetic model. Dom p: p value in a dominant genetic model of inheritance. OR: odds ratio in an allelic genetic model. CI: confidence interval.

|  |  | Control |  |  |  | Case |  |  |  |  |  |  |
| --- | --- | --- | --- | --- | --- | --- | --- | --- | --- | --- | --- | --- |
| locus | Gene | AA | Aa | aa | Total | AA | Aa | aa | Total | Allelic p | Dom p | OR<br>(95% CI) |
| rs842618 | REL | 883 | 231 | 13 | 1075 | 289 | 99 | 7 | 395 | 0.032 | 0.035 | 1.33<br>(1.02 –<br>1.73) |
| rs842634 | REL | 901 | 218 | 11 | 1130 | 299 | 92 | 6 | 397 | 0.052 | 0.064 | 1.21<br>(0.72-2.0) |

### BHLHE40 variants are associated with pediatric TB in South Africa

We next evaluated whether variants in CNBP, REL, and BHLHE40 were associated with pediatric TB in South Africa (Figure E3) (40). BHLHE40 SNP rs11130215 was associated with decreased risk for pediatric TB in an allelic model (Table 2 and Table E5; p = 0.001) _-_ which best fit a dominant model of inheritance p = 3.3×10^−4^, OR 0.5 (0.33 – 0.75). Rs11130215 was in low LD with rs4496464 in the South African cohort (R^2^ 0.10, D’ 0.30). To adjust for ethnic heterogeneity, we genotyped a panel of 95 ancestry informational markers (AIMs) and performed principal components analysis, as described previously (37). The association between rs11130215 and pediatric TB remained statistically significant after adjustment for gender and the top five principal components of the tested AIMs (Table 2, p = 0.01, OR 0.24 - 0.83). No REL or CNBP SNPs were associated with pediatric TB, including rs842634 and rs11709852. Together, these data suggest that a BHLHE40 polymorphism (rs11130215) linked to rs4496464 and increased IL-10 expression is associated with a decreased risk for pediatric TB.

**Table 2.** Association of SNPs with pediatric TB in South Africa. Number of individuals with major homozygous (AA), heterozygous (Aa), and minor homozygous (aa) genotypes described. Allelic p: p value in an allelic genetic model. Dom p: p value in a dominant genetic model by logistic regression with adjustment for ancestry and gender. OR: odds ratio; CI: confidence interval. * adjusted for ethnicity and gender by logistic regression.

| locus | Gene | Control |  |  |  | Case |  |  |  | Allelic p | Dom p | OR (95% CI) |
| --- | --- | --- | --- | --- | --- | --- | --- | --- | --- | --- | --- | --- |
|  |  | AA | Aa | aa | Total | AA | Aa | aa | Total |  |  |  |
| rs11130215 | BHLHE40 | 99 | 169 | 65 | 333 | 78 | 67 | 25 | 170 | 0.001 | 3.3x10<sup>-4</sup> | 0.5 (0.33 – 0.75) |
|  |  |  |  |  |  |  |  |  |  |  | 0.012* | 0.56 (0.28 – 0.87)* |
| rs4496464 | BHLHE40 | 158 | 141 | 35 | 334 | 86 | 66 | 17 | 169 | 0.51 | 0.48 | 1.21 (0.72–2.0) |
|  |  |  |  |  |  |  |  |  |  |  | 0.39* | 1.30 (0.71–2.4)* |

### CREL, CNBP and BHLHE40 SNPs are not associated with BCG-induced T cell responses in South African infants

We next examined whether these variants were associated with adaptive immune responses as a possible mechanism of TB susceptibility due to DC regulation of T cell responses. We tested this hypothesis in a cohort of South African infants that were vaccinated with BCG at birth and whose BCG-specific CD4+ IL-2, TNF, and IFNγ+T cell responses were measured at 10 weeks of age by flow cytometry (36, 37) (Figure E4). Overall media (Figure 7A), BCG-induced (Figure 7B), and SEB-induced (Figure 7C) responses are shown. We evaluated the association between genetic variation in our SNPs of interest: rs842634, rs11709852, rs4496464, and rs11130215, with the frequency of BCG-induced IL-2, TNF, and IFNγ in CD4+ T-cells. Rs11709852 and rs842634 were monoallelic in the South African cohort and not analyzed further. Rs4496464 was associated with a trend toward increased IL2+CD4+ T cell frequency after BCG re-stimulation but this did not achieve statistical significance (Figure 7D, p = 0.15, generalized linear model). This SNP was not associated with TNF or IFNγ frequency in CD4+ T cells (Figure 7E-F). The G allele of BHLHE40 rs11130215 was associated with increased frequency of BCG-specific IL2+CD4+ cells (Figure 7G, p = 0.015, generalized linear model), but not TNF or IFNγ (Figure 7H-I). In a second validation cohort, rs11130215 was associated with a trend toward increased IL-2 expression that did not achieve statistical significance (Figure 7J, p = 0.06, generalized linear model). However, when these data were combined, we found that this SNP was associated with increased IL-2 from CD4+ T cells (Figure 7K, p = 0.006, generalized linear model). Taken together, these data suggest that a BHLHE40 variant is associated with increased IL-2-producing CD4+ T cells, and decreased risk for pediatric TB in a genetic cohort of South African infants.

**Figure 7.**
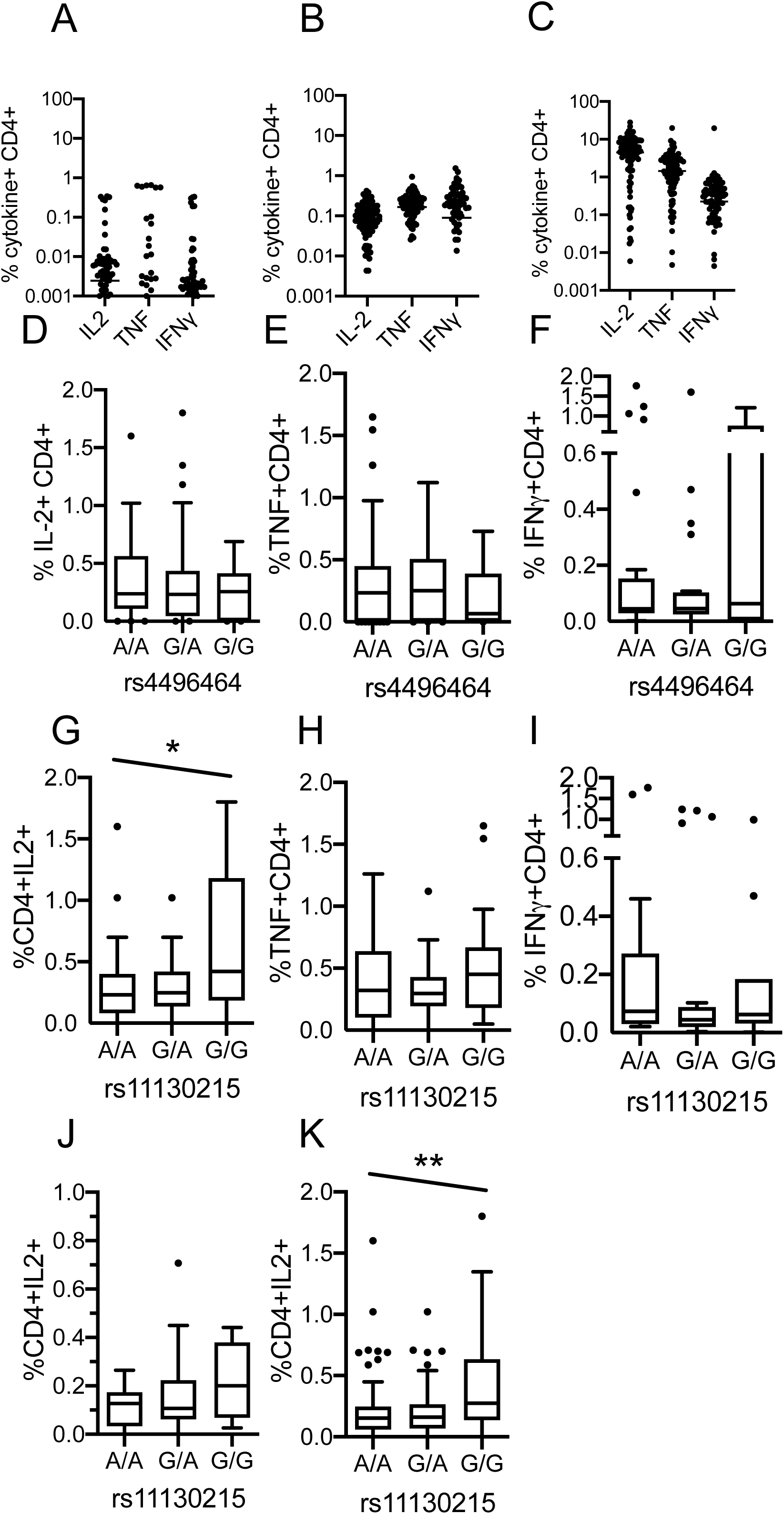
BHLHE40 SNP rs11130215 is associated with BCG-induced IL-2+CD4+ T-cell responses in South African infants. BCG-specific CD4+ T cell responses from South African infants at 10 weeks of age were measured by flow cytometry and stratified by genotype of interest. Background correction was performed by subtracting the proportion of cytokine-producing cells after BCG or SEB stimulation from media control stimulation. A-C) A) Media control, B) BCG-induced, and C) staphylococcus enterotoxin B (SEB)- induced IL-2, TNF, and IFNγ+ CD4+ T cell responses. N = 88. D-F) We measured the frequency of BCG-specific D) IL-2+, E) TNF+, and F) IFNγ+ CD4+ T cells after 12 hours of re-stimulation and stratified by rs4496464. A/A N = 29, G/A N = 44, G/G N = 11. G-I) We measured the frequency of BCG-specific G) IL-2+, H) TNF+, and I) IFNγ+ CD4+ T cells after 12 hours of re-stimulation and stratified by rs11130215 in a discovery cohort. A/A N = 24, G/A N = 31, G/G N = 19. J) Proportion of BCG-specific IL-2+CD4+ T cells, stratified by rs11130215, in an independent validation set. A/A N = 26, G/A N = 47, G/G N = 20. K) Combined datasets from D) and I). All data visualized as Tukey plots, with middle bar representing median, thick bars with interquartile range, and whiskers drawn to 10-90^th^ percentile. Outliers are represented with dots. * p < 0.05, ** p < 0.01, generalized linear model.

## Discussion

IL-12 and IL-10 are both essential for an effective host response to tuberculosis, and overexpression of either cytokine can similarly lead to adverse outcomes. In this paper, we found that variation in REL and BHLHE40, genes that directly influence expression of these cytokines, is associated with secretion of IL-12 and IL-10, respectively, from peripheral blood DCs using a flow cytometry-based assay. To our knowledge, this assay has not been used previously to evaluate the genetics of DC immune responses (20, 41). Related variants in REL were associated with increased expression of IL-12 and also with increased susceptibility to TBM, and SNPs in BHLHE40 associated with increased IL-10 were also associated with decreased risk for pediatric TB. These data represent the most comprehensive evaluation of the human genetic loci associated with IL-10 and IL-12 production in TB pathogenesis.

Both insufficient and excessive IL-10 responses are harmful to TB control (32, 42). We found BHLHE40 variants that were associated with increased IL-10 production in myeloid cells after LPS and TB whole cell lysate stimulation. A variant in linkage disequilibrium was also associated with increased BCG-specific IL-2+CD4+ T cells with stable frequencies of TNF+ and IFNγ+ CD4+ T cells in South African infants. Critically, this variant was associated with decreased risk for developing pediatric TB. Canonically, increased IL-10 is associated with increased differentiation of regulatory T cells (43), which may delay the appropriate activation of effective adaptive immune responses to Mtb (44). However, a balanced immune response with increased number of antigen-specific T cells overall is beneficial to preventing infection. The relatively modest changes to the cytokine response associated with genotype may influence T cell proliferation and differentiation to promote a balanced and effective T cell response (45). Moreover, BHLHE40 also demonstrates direct effects on T cell function in murine models, and may be an alternate mechanism for the phenotypes we observed (46). IL-10 decreases pathology that may promote effective Mtb control (34, 47). Our observations are consistent with a model whereby modest increases in BHLHE40 are associated with increased IL-10 in macrophages, expanded IL-2+CD4+ T cell responses, and protection from TB. Notably, these data support findings from the mouse model, where BHLHE40 deficiency was associated with early Mtb death due to excessive neutrophil-dominant inflammatory response (34). Study of the factors that influence IL-10 expression may provide insight into a suite of macrophage or T cell changes that may provide insight into TB susceptibility and control.

Variation in REL rs842634 was associated with increased IL-12 production from dendritic cells after LPS and TBWCL stimulation. A SNP in linkage disequilibrium, rs842618, was also associated with increased risk for TB meningitis in a Vietnamese cohort. Although IL-12 is canonically associated with protection from TB, significant evidence has accumulated that increases in proinflammatory cytokines, including TNF and IFNγ, may also be harmful for Mtb control in some settings, including TBM (12, 45, 48). Although IL-12α and IFNγ are essential for control of Mtb infection, the amount necessary for protection remains unclear (45). Excessive IFNγ induces immune pathology requiring anti-inflammatory therapy during TB immune reconstitution syndrome (49). IL-12 also induces TNF, in CD4+ T cells as part of the Th1 response (50). Excess TNF in Mtb-infected macrophages leads to necrosis and Mtb spread, and worsens TBM outcomes (51). Identification of genetic factors that modulate dendritic cell proinflammatory cytokines provides insight into the optimal balance of cytokines to control Mtb in adults.

This study has several potential limitations. We do not yet have evidence of functional SNPs that directly regulate gene function. Future fine-mapping studies with in vitro mechanistic assays will be required to determine the specific alleles that regulate cellular function and clinical outcomes together. A second limitation is that some of these observations do not achieve statistical significance after adjustments for multiple comparisons with associations with clinical outcomes. Although this limitation is true for the clinical findings, the evidence supporting a genetic regulatory role of human cellular IL12/IL10 responses was robust and provided support for the possible clinical associations. Given this, we used a threshold of p < 0.05 as a measure of statistical significance, without the conservative Bonferroni correction. Further studies will be needed in additional cohorts, particularly after discovery of the causal SNP that regulates cytokine production. Third, case-control studies of TB outcomes may have misclassification of controls, as we examined population controls in studies in our Vietnamese cohort. However, classification errors that arise from such control populations likely lead to reduction in the statistical power of these studies.

To our knowledge, this study represents the most comprehensive analysis to date of genetic regulation of dendritic cell IL-12 and IL-10 production by common polymorphisms and their association with TB outcomes. Although further studies are required, overlapping genetic studies of immune outcomes and TB clinical susceptibility may lead to important breakthroughs in TB vaccine design and immune drug development.

## Data Availability

All data collected in this manuscript is publicly available upon request to the corresponding author.

## Acknowledgements

The authors thank the individuals and families who participated in the study. They also thank the immunology and clinical teams at the hospitals in Ho Chi Minh City, Vietnam and Worcester, South Africa for obtaining informed consent and collecting and processing samples from study participants. They acknowledge the support of the Center for Emerging and Reemerging Infectious Disease Flow Cytometry Facility at the University of Washington.

## Notes

### Competing Interest Statement

The authors have declared no competing interest.

### Funding Statement

This work was supported by funding from the following NIH grants: R01 AI136921 to JAS; P01 AI132130 to CMS, TRH, JAS, SJD, TJS; K24 AI137310 to TRH; AI067497 to KAF. No authors received any other payment or services from a third party for any aspect of the submitted work.

