## Supplementary material for "CNBP, REL, and BHLHE40 variants are associated with IL-12 and IL-10 responses and tuberculosis risk": Online Supplement

**Title: CNBP, REL, and BHLHE40 variation is associated with IL12 and IL10 responses and tuberculosis outcomes.**

**Online Supplement**

**Supplemental Materials and Methods**

*Reagents and Antibodies*

RPMI 1640 medium, L-glutamate were obtained from Life Technologies. Ultrapure LPS (TLR4 ligand) isolated from *Salmonella Minnesota* R595 was obtained from List Biological Labs. Whole cell lysate from *M. tuberculosis* strain H37Rv was obtained as part of National Institutes of Health, National Institute of Allergy and Infectious Diseases Contract No. HHSN266200400091C, entitled Tuberculosis Vaccine Testing and Research Materials (Colorado State University, Fort Collins, CO). Lyophilized live Bacille Calmette-Guerin (BCG, 20 x 10^6^ CFU/mL) was obtained from Statens Serum Institute (Copenhagen, Denmark).

Antibodies (clones and source) for flow cytometry with intracellular cytokine staining are shown in **Table E1 (Key Resources Table)**. The concentrations of all antibodies were titrated prior to use.

*Study Participants and Ethics Statement*

Approval for human study protocols was obtained from the institutional review boards at local sites in Vietnam, South Africa, and the University of Washington School of Medicine. Genomic DNA was purified from blood samples using genomic DNA isolation kits (Qiagen, Inc). DNA concentrations were confirmed using Nanodrop. For the Seattle cohort, study subjects were local volunteers self-described as healthy without history of recurrent of serious infections. 52% of individuals are female, and 48% are male. The ethnic composition of this study group was 69% White, 19% Asian, 2% Black or African American, and 2% Latinx. Average age of study participants was 39, with interquartile range of 29 – 46 at the time of their enrollment.

For genetic studies in Vietnam, approval for human study protocols was obtained from the human review boards at the University of Washington School of Medicine, the Hospital for Tropical Diseases (Ho Chi Minh City, Vietnam), Pham Ngoc Thach Hospital for Tuberculosis and Lung Diseases (Ho Chi Minh City, Vietnam), Health Services of Ho Chi Minh City, Hung Vuong Hospital (Ho Chi Minh City, Vietnam), and the Oxford Tropical Research Ethics Committee. For South African pediatric TB cohorts, the study was conducted according to the U.S. Department of Health and Human Services and Good Clinical Practice guidelines. This study included written informed consent from the parent or legal guardian of the study participant and protocol approval by the University of Cape Town Research Ethics Committee and the University of Washington human subjects review board.

South African study participants were enrolled at the South African Tuberculosis Vaccine Initiative field site in Worcester, South Africa, near Cape Town as part of a larger study on BCG vaccination with 11,680 infants (1, 2). This area has one of the highest rates of TB incidence in the world with an incidence of 3% among children under 3 years of age in the study population (1, 2). A nested genetics case-control study was performed with identification of cases and controls during a 2-year prospective observation period after vaccination at birth. Household controls were children without TB disease over the 2-year follow period living with an individual with active TB disease. Community controls had no history of TB disease in the first two years of life. Community-wide passive surveillance systems identified patients with TB disease and children with symptoms suggestive of TB disease. The criteria for detection of TB cases have been described previously (3). All infants who had symptoms compatible with TB disease or who had contact with an adult with TB disease were admitted to a dedicated research ward for clinical examination, chest radiography, tuberculin skin testing, two early-morning gastric aspirations, and two sputum inductions for Mtb smear and culture. Participants were defined as having “definite TB” if they had a positive Mtb culture, a positive smear, or a positive Mtb PCR from one of their samples. Participants with a chest radiograph compatible with or suggestive of TB combined with one or more additional laboratory or clinical features consistent with TB (smear negative, cough > 2 weeks, PPD skin test ≥ 15 mm, failure to thrive, and recent weight loss) were defined as having “probable TB.” Individuals without radiography consistent with TB who were diagnosed with TB by the treating physician and had 2 or more clinical features suggestive of TB were defined as “possible TB.” All others were described as “not TB.” All infants admitted to the research ward were also tested for HIV infection and positive tests resulted in exclusion from the study. The following were additional exclusion criteria at 10 weeks of age: mother known to be infected with HIV; BCG not received by infant within 24 hours of birth; significant perinatal complications in the infant; any acute or chronic disease in the infant at the time of enrollment; clinically apparent anemia in the infant; household contact with any person with TB disease or any person who was coughing.

Study subjects from the Vietnam cohort were described previously and are briefly

summarized here (4). Subjects with tuberculous meningitis (TBM) were recruited from two centers in Ho Chi Minh City, Vietnam: Pham Ngoc Thach (PNT) Hospital for Tuberculosis and the Hospital for Tropical Diseases (HTD). Subjects with pulmonary TB were recruited from a network of district TB control units within Ho Chi Minh City that provide directly observed therapy to TB patients. In addition, pulmonary TB subjects were recruited from PNT hospital from 2006 through 2008. Vietnamese population controls are otherwise healthy adults with primary angle closure glaucoma which have been previously described (5). All case and control participants were unrelated, and greater than 95% were of the Vietnamese Kinh ethnicity. Previous genetic studies of this population indicate minimal population substructure (4). Written, informed consent was obtained from patients or their relatives if the patient could not provide consent (i.e., was unconscious). Individuals in the TBM group were defined as follows. Individuals at least 15 years old, admitted to these centers with clinical meningitis (defined as nuchal rigidity and abnormal cerebrospinal fluid parameters), a negative HIV test result, and a positive Ziehl–Neelsen stain for acid-fast bacilli or Mtb culture, or both, from cerebrospinal fluid (“definite TBM”) were recruited for genetics studies from 2001 to 2008. In addition to definite TBM, the cohort included subjects with “probable TBM,” defined as clinical meningitis plus at least one of the following: chest radiograph consistent with active TB, acid-fast bacilli found in any specimen other than cerebrospinal fluid, and clinical evidence of other extrapulmonary TB. The pulmonary TB group was defined as follows: participatns were outpatients who were at least 18 years old, had no previous history of treatment for TB, no evidence of miliary or extrapulmonary TB, chest radiograph results consistent with non-miliary pulmonary TB, negative HIV test results, and sputum smear positive for acid-fast bacilli or M. tuberculosis cultured from sputum.

*Stimulation of whole blood samples, cell culture, and flow cytometry*

LPS (10 ng/ml), BCG (MOI=5), and TB whole cell lysate (TBWCL; 50 µg/ml) were added to 500 µl of whole blood for predetermined times. Media-stimulated cells were included as a negative control. Brefeldin A (BFA, Sigma) was added at a concentration of 500 μg/mL (50 times the desired final concentration of 10 ng/mL) additionally Protein Transport Inhibitor containing monensin (monensin, BD) was added according to manufacturer’s protocol to all wells 4 hours prior to the completion of the experiment. Primary monocytes were isolated from human subjects as described previously (6). Briefly, peripheral blood was obtained and PBMCs were collected via Ficoll gradient separation and cryopreserved. Subsequently, CD14+ cells were purified using human monocyte negative selection kit (Miltenyi Biotec, Inc.). Monocytes were >95% pure for CD14+ using this method.

Infant T cell assays were collected as described previously (3) (7). At 10 weeks of age, heparinized blood was collected from BCG- vaccinated infants and 1 mL was incubated ex vivo with 1.26106 CFU of BCG (Danish strain 1331). None of the infants in this study had active tuberculosis at the time of their 10 week blood draw or during 2 years of follow-up observation. Whole blood was incubated for 12 hours with either media control , BCG, or SEB, then fixed and frozen in liquid nitrogen.

Frozen samples were thawed in a 37°C water bath and spun, then pellets were resuspended in 200 μl PermWash Solution (BD) and incubated at room temperature for 10 min. After one wash in PermWash, cells were stained for 30 min at room temperature. After two further washes with PermWash, cells were immediately analyzed on an LSRII Flow Cytometer (BD). Positivity thresholds were determined by gating on flow-minus-one controls followed by selective testing of thresholds on negative controls and stimulated controls.

*Genotyping and linkage disequilibrium*

Genotyping was performed with Illumina MegaEx Chip for the Seattle and South Africa cohorts and Illumina OmniExpress in Vietnam. Imputation in Vietnam was performed as described previously (8). Selected genotyping was also performed using a Fluidigm Genotyping 96 x 96 array. We selected a region 10 kb upstream and downstream of genes of interest using a minor allele frequency cut-off of 5%. SNPs were excluded if they demonstrated Hardy-Weinberg equilibrium (HWE) p<0.001. Genotypes were determined by Illumina MEGAex array annotated to the human genome (hg19) with annotatr basic genes (v1.14.0) in R (v4.0.2) (9). Linkage disequilibrium (LD) Pearson coefficient of correlation (R^2^) was calculated between SNPs with two or more genotypes and within 10 kb of a gene using the genetics package (v1.3.8.1.2).

*Statistical methods*

All analyzed SNPs were tested for Hardy-Weinberg equilibrium in control subjects using a χ^2^ goodness-of-fit test and were excluded if p<0.001. In our primary analysis, we examined whether polymorphism genotype frequencies were associated with cytokine production using a generalized linear model. For secondary analyses, SNPs were investigated for associations under additional genetic models (dominant, recessive, and additive). In the recessive model, carriers of allele 1 (00 and 01 genotypes) were compared with homozygous subjects for allele 2 (11 genotype). In the dominant model, carriers of allele 2 (01 and 11 genotypes) were compared with homozygous subjects for allele 1 (00 genotype). Graphs were created with Prism version 8.0 (GraphPad, Inc.). Results are reported without correction for multiple comparisons due to the heterogeneous sources of data (mixture of cellular and clinical), including varied availability of validation datasets for some datasets.

**Supplemental Data**

**Figure E1. Genomic Maps and Linkage Disequilibrium Plots for Seattle Cohort**

A-C) R^2^ LD among SNPs evaluated in the Seattle cohort (N = 84) for A) REL, B) CNBP, and C) BHLHE40 gene regions. LD plot depicts SNPs with minor allele frequency (MAF) > 0.1 with depiction of R^2^ LD values (higher R^2^ value with deeper shade of red). Duplicate SNPS (R^2^ LD = 1.0) were merged. Green bars indicate graphical representation of MAF.

D-F) Genomic map depicts SNP base pair positions from Genome Reference Consortium assembly GRCh37 for D) REL (genomic region depicted spans base pair ch2:60,840K – 60,936K), E) CNBP (spans base pair ch3:129,115K -129,192K), and F) BHLHE40 (spans base pair 4,895K – 4,986K).

**Figure E2. Genomic Maps and Linkage Disequilibrium Plots for Vietnam Cohort**

A-C) Linkage disequilibrium (LD) R^2^ values among SNPs evaluated in the Kinh Vietnamese population by 1000 genomes data cohort (N = 102) for A) REL, B) CNBP, and C) BHLHE40 gene regions. LD plot depicts SNPs with minor allele frequency (MAF) > 0.1 with depiction of R^2^ LD values (higher R^2^ value with deeper shade of red). Duplicate SNPS (R^2^ LD = 1.0) were merged. Green bars indicate graphical representation of MAF.

D-F) Genomic map depicts SNP base pair positions from Genome Reference Consortium assembly GRCh37 for D) REL (genomic region depicted spans base pair ch2:60,840K – 60,936K), E) CNBP (spans base pair ch3:129,115K -129,192K), and F) BHLHE40 (spans base pair 4,895K – 4,986K).

**Figure E3: Genomic Maps and Linkage Disequilibrium Plots for South Africa Cohort**

A-C) R^2^ LD among SNPs evaluated in the South African cohort healthy controls (N = 435) for A) REL, B) CNBP, and C) BHLHE40 gene regions. LD plot depicts SNPs with minor allele frequency (MAF) > 0.1 with depiction of R^2^ LD values (higher R^2^ value with deeper shade of red). Duplicate SNPS (R^2^ LD = 1.0) were merged. Green bars indicate graphical representation of MAF.

D-F) Genomic map depicts SNP base pair positions from Genome Reference Consortium assembly GRCh37 for D) REL (genomic region depicted spans base pair ch2:60,840K – 60,936K), E) CNBP (spans base pair ch3:129,115K -129,192K), and F) BHLHE40 (spans base pair 4,895K – 4,986K).

**Figure E4. Gating strategy for South African infant BCG immune response cohort.** Whole blood was obtained from 10-week-old South African infants, stimulated with media, BCG, or SEB for 12 hours, then fixed and frozen. Samples were thawed and stained with fluorescent antibodies. Singlets were selected by FSC-A/FSC-H ratio, then CD14+ cells were excluded and CD3+ cells included. CD4+ cells were selected and the proportion of IL-2+, TNF+, and IFNγ+ T cells were determined.

**Table E1. Key Resources**

| **Panel** |  |  |  |  |
| --- | --- | --- | --- | --- |
| **Antibody** | **Clone** | **Fluorophore** | **Manufacturer** | **Catalog #** |
| CD3 | UCHT1 | PE-Tx Red | Beckman Coulter | IM2705U |
| CD11c | S-HCL-3 | APC | BD | 340714 |
| CD14 | M5E2 | V500 | BD | 561391 |
| CD16 | 3G8 | BV650 | Biolegend | 302042 |
| CD40 | 5C3 | BUV395 | BD | 565202 |
| CD66 | ASL-32 | Biotin | Biolegend | custom |
| CD123 | 6H6 | PE-Cy7 | Biolegend | 306010 |
| HLADR | LN3 | SB600 | eBioscience | 63-9956-42 |
| IFNα | 7N4-1 | PE | BD | 560097 |
| IFNγ | 4S.B3 | BV711 | Biolegend | 502540 |
| IL6 | MQ213A5 | PerCP-EF710 | eBioscience | 46-7069-42 |
| IL10 | BT-10 | FITC | eBioscience | BMS131-2FI |
| IL12 | C8.6 | EF450 | eBioscience | 48-7129-42 |
| Streptavidin |  | BV786 | BD | 563858 |
| TNF | Mab11 | Ax700 | BD | 557996 |
| AVID | Near IR | APC-H7 | Thermofisher | L34976 |

**Table E2**. P values, by generalized linear model (GLM), between SNP genotypes in REL, CNBP, and BHLHE40 gene regions and IL-10 and IL-12 responses from peripheral blood DC in the Seattle healthy donor cohort after 24 hours of stimulation with TB whole cell lysate (TBWCL; 50 μg/ml) or LPS (10 ng/ml).

| SNP | Gene | TBWCL IL10 | TBWCL IL12 | LPS IL10 | LPS IL12 |
| --- | --- | --- | --- | --- | --- |
| rs11709852 | CNBP | 0.3693 | 0.0025 | 0.9081 | 0.9874 |
| rs73212424 | CNBP | 0.3584 | 0.9515 | 0.0273 | 0.6922 |
| rs79248174 | CNBP | 0.0004 | 0.3259 | 0.4303 | 0.7801 |
| rs58457030 | CNBP | 0.7439 | 0.6702 | 0.2542 | 0.7842 |
| rs1177206 | REL | 0.1336 | 0.9426 | 0.1538 | 0.8314 |
| rs1177211 | REL | 0.1622 | 0.9727 | 0.2056 | 0.5000 |
| rs2901182 | REL | 0.7433 | 0.6137 | 0.6057 | 0.3977 |
| rs34695944 | REL | 0.6392 | 0.7571 | 0.2455 | 0.5364 |
| rs3732179 | REL | 0.2658 | 0.2497 | 0.0141 | 0.9767 |
| rs842634 | REL | 0.4555 | 0.0339 | 0.1992 | 0.0439 |
| rs13085159 | BHLHE40 | 0.5058 | 0.3437 | 0.6124 | 0.3703 |
| rs13092419 | BHLHE40 | 0.4170 | 0.4646 | 0.7838 | 0.6056 |
| rs1514453 | BHLHE40 | 0.6247 | 0.7911 | 0.4926 | 0.1854 |
| rs17041710 | BHLHE40 | 0.7512 | 0.4960 | 0.6248 | 0.8312 |
| rs17041758 | BHLHE40 | 0.6422 | 0.4362 | 0.8463 | 0.3463 |
| rs1955179 | BHLHE40 | 0.5633 | 0.7153 | 0.6301 | 0.3731 |
| rs4496464 | BHLHE40 | 0.0052 | 0.1504 | 0.1756 | 0.4715 |
| rs7640124 | BHLHE40 | 0.7771 | 0.1208 | 0.4145 | 0.4319 |
| rs6442917 | BHLHE40 | 0.4017 | 0.8605 | 0.2612 | 0.3874 |
| rs6442925 | BHLHE40 | 0.3163 | 0.3265 | 0.4052 | 0.0962 |
| rs6777136 | BHLHE40 | 0.8002 | 0.4664 | 0.6109 | 0.5178 |
| rs6808127 | BHLHE40 | 0.3909 | 0.4536 | 0.3590 | 0.8543 |
| rs728389 | BHLHE40 | 0.9136 | 0.8018 | 0.3044 | 0.9563 |
| rs7628494 | BHLHE40 | 0.9423 | 0.8382 | 0.0565 | 0.3190 |
| rs7635516 | BHLHE40 | 0.2403 | 0.3156 | 0.6455 | 0.3726 |
| rs7638383 | BHLHE40 | 0.7569 | 0.3436 | 0.1827 | 0.9937 |
| rs9836120 | BHLHE40 | 0.8172 | 0.8298 | 0.5425 | 0.2847 |
| rs34892180 | BHLHE40 | 0.8002 | 0.4664 | 0.6109 | 0.5178 |
| rs7652330 | BHLHE40 | 0.8907 | 0.2090 | 0.6163 | 0.1742 |
| rs11130215 | BHLHE40 | 0.4384 | 0.2995 | 0.2312 | 0.2076 |

**Table E3. Genetic Association between SNPs in CNBP, REL, and BHLHE40 and TB meningitis in Vietnam.** SNP rs ID number depicted along with the common allele (A1) and the uncommon allele (A2), frequency of A2 among cases (F_case) and controls (F_control), and the Chi Square, P values and odds ratio (OR) in an allelic genetic model. Excel file attached.

**Table E4. Genetic Association between SNPs in CNBP, REL, and BHLHE40 and pulmonary TB in Vietnam.** SNP rs ID number depicted along with the common allele (A1) and the uncommon allele (A2), frequency of A2 among cases (F_case) and controls (F_control), and the Chi Square, P values and odds ratio (OR) in an allelic genetic model. Excel file attached.

**Table E5. Genetic Association between SNPs in CNBP, REL, and BHLHE40 and pediatric TB in South Africa**. SNP rs ID number depicted along with the common allele (A1) and the uncommon allele (A2), frequency of A2 among cases (F_case) and controls (F_control), and the Chi Square, P values and odds ratio (OR) in an allelic genetic model. Excel file attached.

5. Khor CC, Do T, Jia H, Nakano M, George R, Abu-Amero K, Duvesh R, Chen LJ, Li Z, Nongpiur ME, Perera SA, Qiao C, Wong HT, Sakai H, Barbosa de Melo M, Lee MC, Chan AS, Azhany Y, Dao TL, Ikeda Y, Perez-Grossmann RA, Zarnowski T, Day AC, Jonas JB, Tam PO, Tran TA, Ayub H, Akhtar F, Micheal S, Chew PT, Aljasim LA, Dada T, Luu TT, Awadalla MS, Kitnarong N, Wanichwecharungruang B, Aung YY, Mohamed-Noor J, Vijayan S, Sarangapani S, Husain R, Jap A, Baskaran M, Goh D, Su DH, Wang H, Yong VK, Yip LW, Trinh TB, Makornwattana M, Nguyen TT, Leuenberger EU, Park KH, Wiyogo WA, Kumar RS, Tello C, Kurimoto Y, Thapa SS, Pathanapitoon K, Salmon JF, Sohn YH, Fea A, Ozaki M, Lai JS, Tantisevi V, Khaing CC, Mizoguchi T, Nakano S, Kim CY, Tang G, Fan S, Wu R, Meng H, Nguyen TT, Tran TD, Ueno M, Martinez JM, Ramli N, Aung YM, Reyes RD, Vernon SA, Fang SK, Xie Z, Chen XY, Foo JN, Sim KS, Wong TT, Quek DT, Venkatesh R, Kavitha S, Krishnadas SR, Soumittra N, Shantha B, Lim BA, Ogle J, de Vasconcellos JP, Costa VP, Abe RY, de Souza BB, Sng CC, Aquino MC, Kosior-Jarecka E, Fong GB, Tamanaja VC, Fujita R, Jiang Y, Waseem N, Low S, Pham HN, Al-Shahwan S, Craven ER, Khan MI, Dada R, Mohanty K, Faiq MA, Hewitt AW, Burdon KP, Gan EH, Prutthipongsit A, Patthanathamrongkasem T, Catacutan MA, Felarca IR, Liao CS, Rusmayani E, Istiantoro VW, Consolandi G, Pignata G, Lavia C, Rojanapongpun P, Mangkornkanokpong L, Chansangpetch S, Chan JC, Choy BN, Shum JW, Than HM, Oo KT, Han AT, Yong VH, Ng XY, Goh SR, Chong YF, Hibberd ML, Seielstad M, Png E, Dunstan SJ, Chau NV, Bei J, Zeng YX, Karkey A, Basnyat B, Pasutto F, Paoli D, Frezzotti P, Wang JJ, Mitchell P, Fingert JH, Allingham RR, Hauser MA, Lim ST, Chew SH, Ebstein RP, Sakuntabhai A, Park KH, Ahn J, Boland G, Snippe H, Stead R, Quino R, Zaw SN, Lukasik U, Shetty R, Zahari M, Bae HW, Oo NL, Kubota T, Manassakorn A, Ho WL, Dallorto L, Hwang YH, Kiire CA, Kuroda M, Djamal ZE, Peregrino JI, Ghosh A, Jeoung JW, Hoan TS, Srisamran N, Sandragasu T, Set SH, Doan VH, Bhattacharya SS, Ho CL, Tan DT, Sihota R, Loon SC, Mori K, Kinoshita S, Hollander AI, Qamar R, Wang YX, Teo YY, Tai ES, Hartleben-Matkin C, Lozano-Giral D, Saw SM, Cheng CY, Zenteno JC, Pang CP, Bui HT, Hee O, Craig JE, Edward DP, Yonahara M, Neto JM, Guevara-Fujita ML, Xu L, Ritch R, Liza-Sharmini AT, Wong TY, Al-Obeidan S, Do NH, Sundaresan P, Tham CC, Foster PJ, Vijaya L, Tashiro K, Vithana EN, Wang N, Aung T. Genome-wide association study identifies five new susceptibility loci for primary angle closure glaucoma. *Nat Genet* 2016; 48: 556-562.

6. Shah JA, Vary JC, Chau TT, Bang ND, Yen NT, Farrar JJ, Dunstan SJ, Hawn TR. Human TOLLIP Regulates TLR2 and TLR4 Signaling and Its Polymorphisms Are Associated with Susceptibility to Tuberculosis. *J Immunol* 2012; 189: 1737-1746.

7. Randhawa AK, Shey MS, Keyser A, Peixoto B, Wells RD, de Kock M, Lerumo L, Hughes J, Hussey G, Hawkridge A, Kaplan G, Hanekom WA, Hawn TR. Association of human TLR1 and TLR6 deficiency with altered immune responses to BCG vaccination in South African infants. *PLoS Pathog* 2011; 7: e1002174.

8. Dunstan SJ, Hue NT, Han B, Li Z, Tram TT, Sim KS, Parry CM, Chinh NT, Vinh H, Lan NP, Thieu NT, Vinh PV, Koirala S, Dongol S, Arjyal A, Karkey A, Shilpakar O, Dolecek C, Foo JN, Phuong le T, Lanh MN, Do T, Aung T, Hon DN, Teo YY, Hibberd ML, Anders KL, Okada Y, Raychaudhuri S, Simmons CP, Baker S, de Bakker PI, Basnyat B, Hien TT, Farrar JJ, Khor CC. Variation at HLA-DRB1 is associated with resistance to enteric fever. *Nat Genet* 2014; 46: 1333-1336.

9. Cavalcante RG, Sartor MA. annotatr: genomic regions in context. *Bioinformatics* 2017; 33: 2381-2383.
