## Supplemental Figures for "CNBP, REL, and BHLHE40 variants are associated with IL-12 and IL-10 responses and tuberculosis risk"

### Slide 1
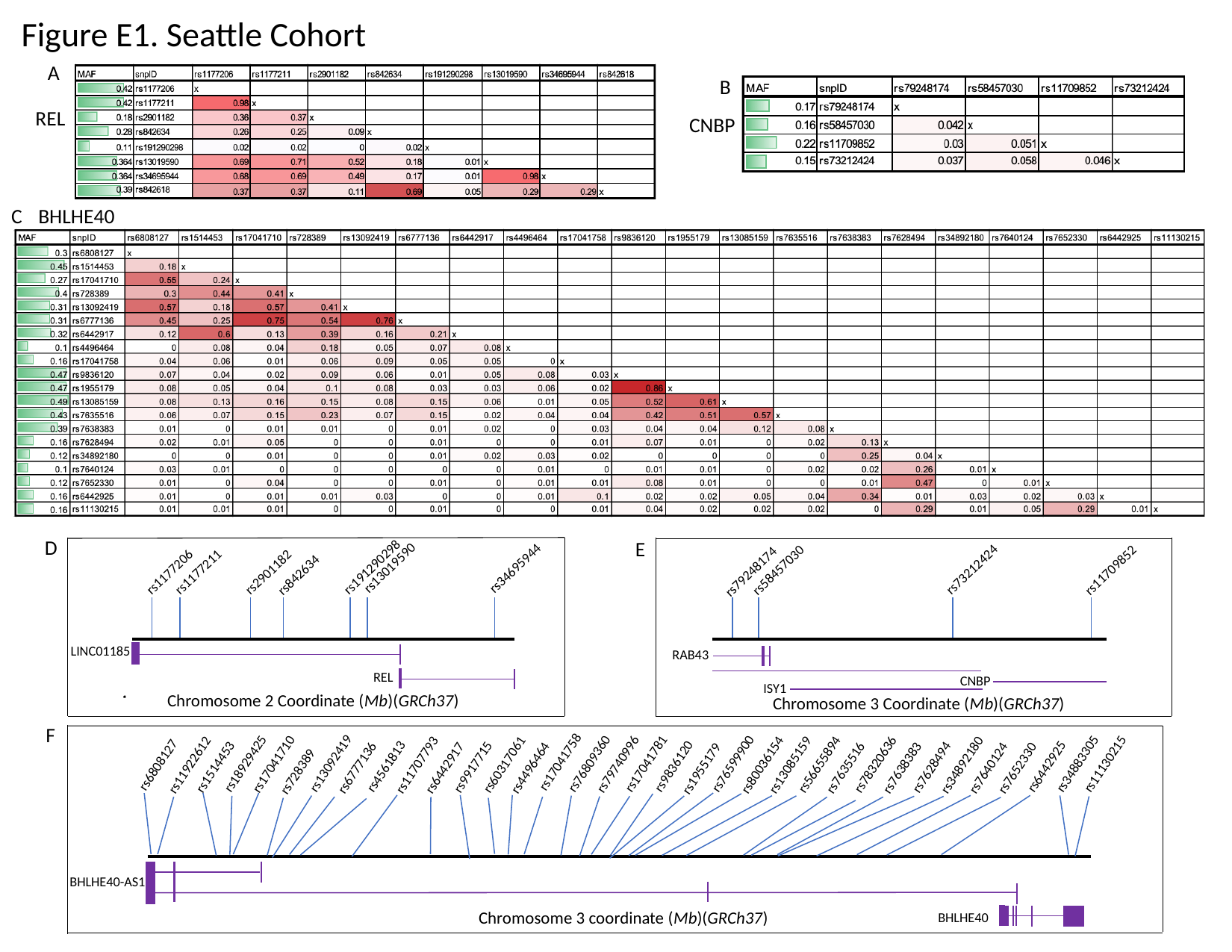

Figure E1. Seattle Cohort
A
B
REL
CNBP
C
BHLHE40
D
E
rs191290298
rs13019590
rs34695944
rs73212424
rs58457030
rs11709852
rs79248174
rs1177211
rs1177206
rs2901182
rs842634
LINC01185
RAB43
REL
CNBP
ISY1
Chromosome 2 Coordinate (Mb)(GRCh37)
Chromosome 3 Coordinate (Mb)(GRCh37)
F
rs17041758
rs13092419
rs18929425
rs76809360
rs17041781
rs76599900
rs34883305
rs11130215
rs17041710
rs79740996
rs78320636
rs56655894
rs60317061
rs11922612
rs11707793
rs80036154
rs13085159
rs34892180
rs6808127
rs9836120
rs4561813
rs6442925
rs1514453
rs9917715
rs7628494
rs1955179
rs6777136
rs4496464
rs7635516
rs7638383
rs7640124
rs6442917
rs7652330
rs728389
BHLHE40-AS1
Chromosome 3 coordinate (Mb)(GRCh37)
BHLHE40

### Slide 2
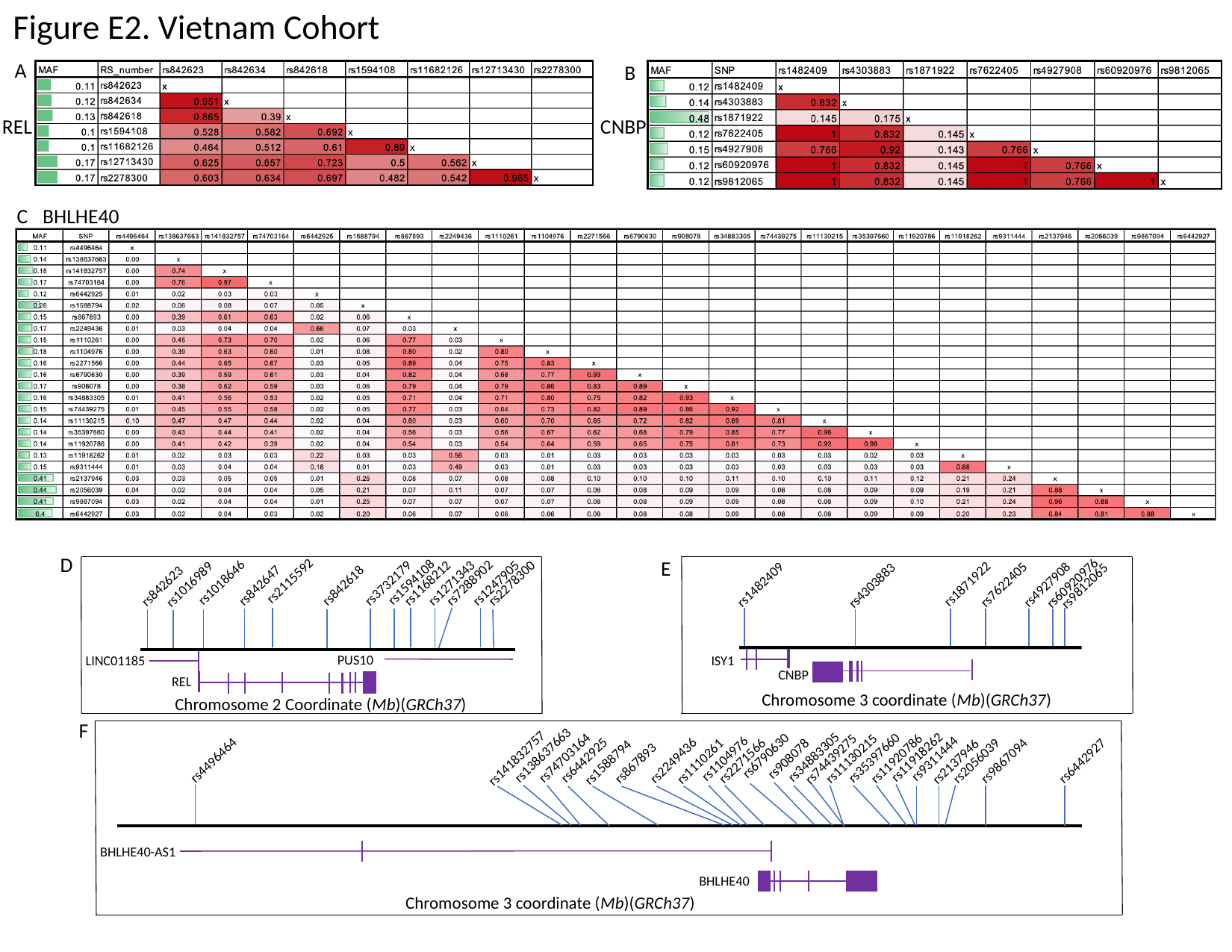

Figure E2. Vietnam Cohort
A
B
REL
CNBP
C
BHLHE40
D
E
rs2115592
rs1594108
rs1018646
rs7288902
rs1168212
rs1271343
rs2278300
rs60920976
rs1247905
rs3732179
rs1871922
rs1016989
rs7622405
rs4927908
rs1482409
rs842647
rs4303883
rs842618
rs9812065
rs842623
PUS10
ISY1
LINC01185
CNBP
REL
Chromosome 3 coordinate (Mb)(GRCh37)
Chromosome 2 Coordinate (Mb)(GRCh37)
F
rs138637663
rs6790630
rs11918262
rs34883305
rs74703164
rs35397660
rs141832757
rs11130215
rs11920786
rs908078
rs74439275
rs9311444
rs1104976
rs4496464
rs6442925
rs2249436
rs2271566
rs9867094
rs2056039
rs6442927
rs1110261
rs2137946
rs1588794
rs867893
BHLHE40-AS1
BHLHE40
Chromosome 3 coordinate (Mb)(GRCh37)

### Slide 3
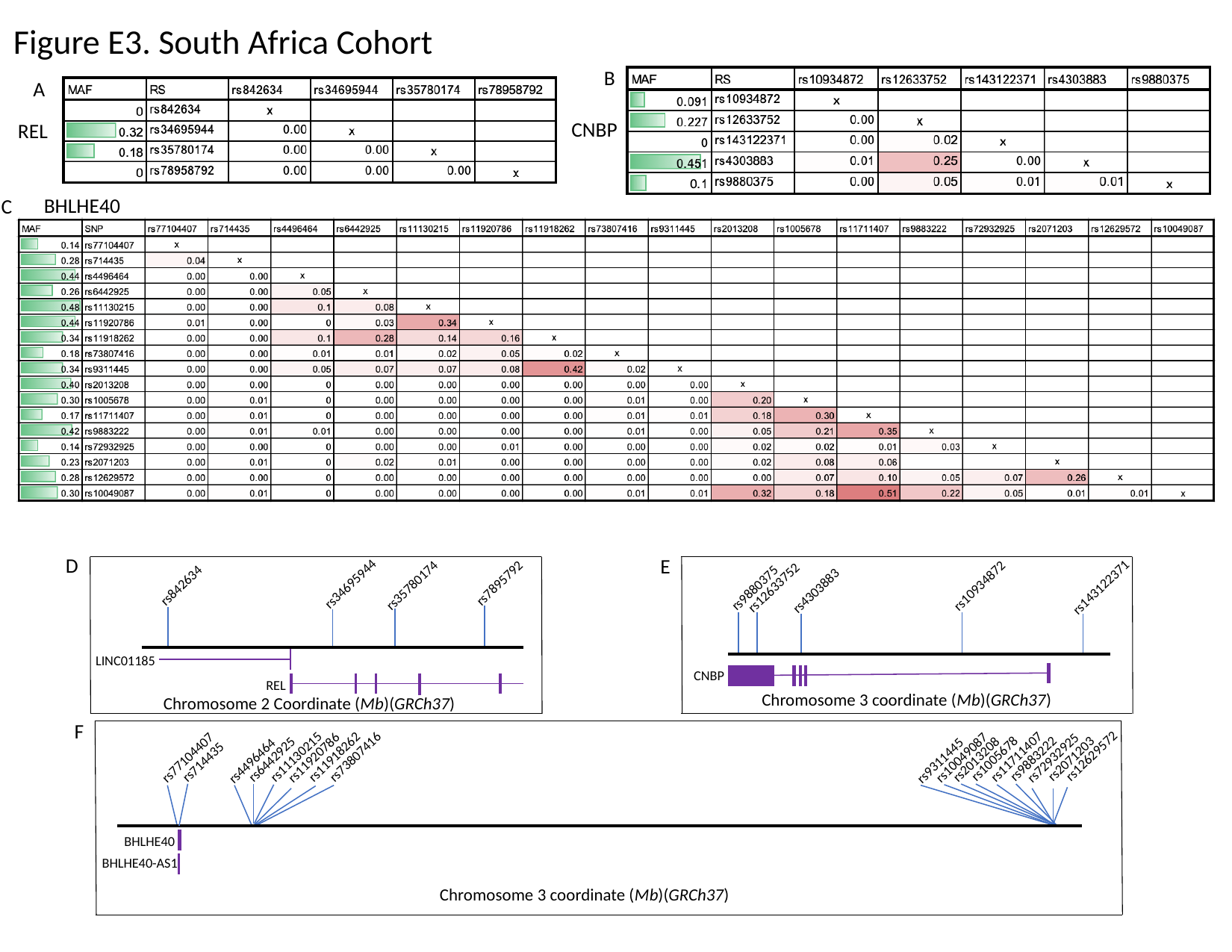

Figure E3. South Africa Cohort
B
A
CNBP
REL
BHLHE40
C
D
E
rs7895792
rs34695944
rs35780174
rs842634
rs10934872
rs143122371
rs12633752
rs9880375
rs4303883
LINC01185
CNBP
REL
Chromosome 3 coordinate (Mb)(GRCh37)
Chromosome 2 Coordinate (Mb)(GRCh37)
F
rs12629572
rs11711407
rs11130215
rs11918262
rs73807416
rs77104407
rs11920786
rs10049087
rs72932925
rs1005678
rs9883222
rs2071203
rs6442925
rs2013208
rs9311445
rs4496464
rs714435
BHLHE40
BHLHE40-AS1
Chromosome 3 coordinate (Mb)(GRCh37)

### Slide 4
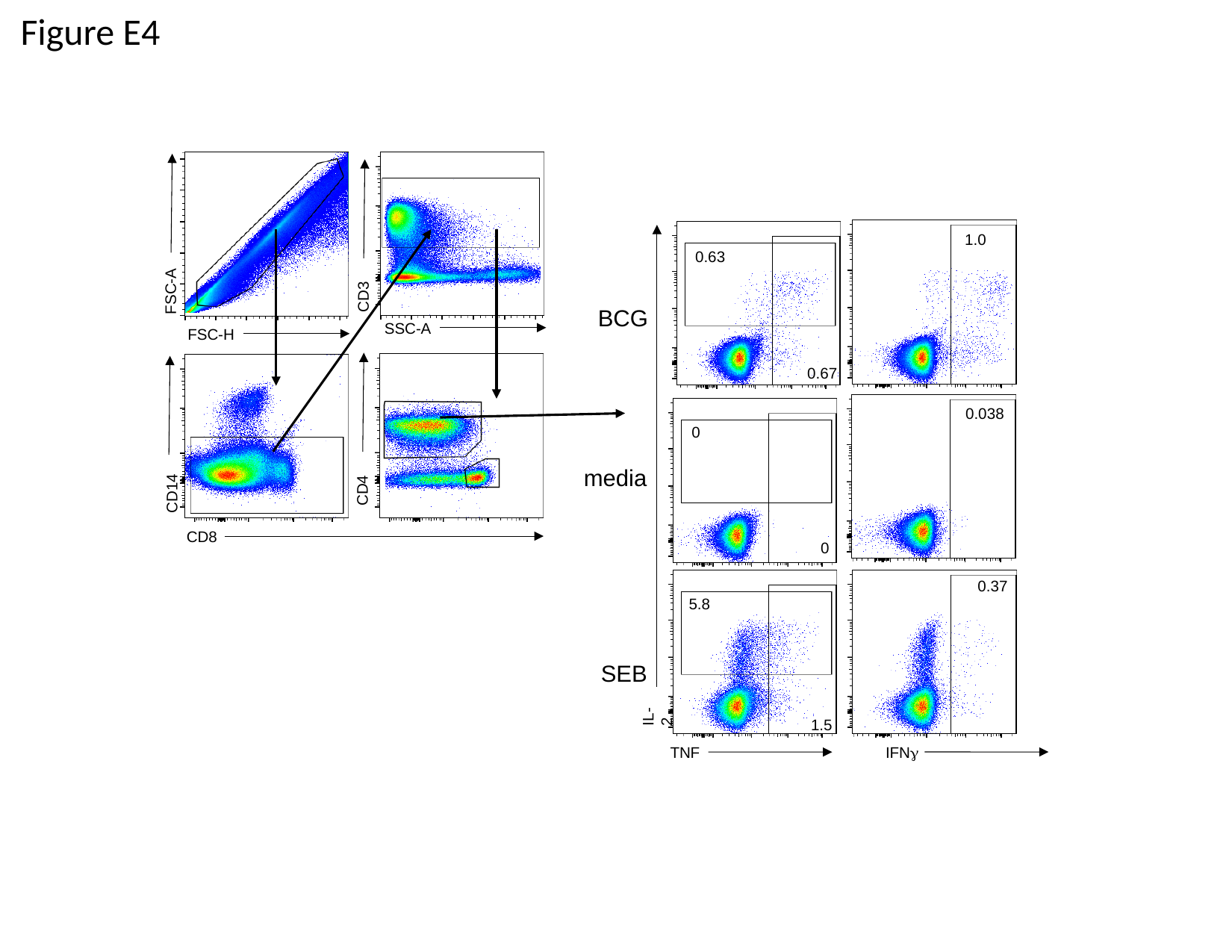

Figure E4
1.0
0.63
FSC-A
CD3
BCG
SSC-A
FSC-H
0.67
0.038
0
media
CD4
CD14
CD8
0
0.37
5.8
SEB
IL-2
1.5
IFNγ
TNF
